# Suicide prediction with natural language processing of electronic health records

**DOI:** 10.1101/2023.09.28.23296268

**Authors:** Alexandra Korda, Marco Heide, Alena Nag, Valerie-Noelle Trulley, Helena- Victoria Rogg, Mihai Avram, Sofia Eickhoff, Kamila Jauch-Chara, Kai Wehkamp, Xingyi Song, Thomas Martinetz, Jörn Conell, Angus Roberts, Robert Stewart, Christina Andreou, Stefan Borgwardt

## Abstract

Suicide attempts are one of the most challenging psychiatric outcomes and have great importance in clinical practice. However, they remain difficult to detect in a standardised way to assist prevention because assessment is mostly qualitative and often subjective. As digital documentation is increasingly used in the medical field, Electronic Health Records (EHRs) have become a source of information that can be used for prevention purposes, containing codified data, structured data, and unstructured free text. This study aims to provide a quantitative approach to suicidality detection using EHRs, employing natural language processing techniques in combination with deep learning artificial intelligence methods to create an algorithm intended for use with medical documentation in German. Using psychiatric medical files from in-patient psychiatric hospitalisations between 2013 and 2021, free text reports will be transformed into structured embeddings using a German trained adaptation of Word2Vec, followed by a Long-Short Term Memory (LSTM) – Convolutional Neural Network (CNN) approach on sentences of interest. Text outside the sentences of interest will be analysed as context using a fixed size ordinally-forgetting encoding (FOFE) before combining these findings with the LSTM-CNN results in order to label suicide related content. This study will offer promising ways for automated early detection of suicide attempts and therefore holds opportunities for mental health care.

## Introduction

Suicide, to this day, remains a significant health risk worldwide, accounting for approximately 700,000 deaths each year in addition to an indeterminably higher number of attempts^1^. In 2019, in Germany alone, on average 25 people took their own life daily, highlighting the undeniable severity of this issue^2^. In tackling the challenge of reducing suicide deaths, the World Health Organization has stressed^1^, among others, the importance of early identification of at-risk individuals as a means of prevention.

Mental health professionals currently assess risk of suicide by analysing the behaviour, language and speech of their patients subjectively in addition to a limited number of standardised scores, a combination of methods that may be improved upon through the usage of artificial intelligence. Electronic Health Records (EHRs) are the most obvious source of structured and unstructured digital information in this context, as they include real-world clinical information (e.g., diagnoses, treatment plans, prescriptions). Importantly, EHRs are being increasingly adopted across healthcare systems internationally, and there have been some approaches to using coded data within EHRs for prediction of psychosis risk^3^ and of mental health outcomes such as hospital re-admissions^4^ or suicide risk^5^.

A challenge in predicting suicide risk based on EHR data is that risk factors are highly heterogeneous and their appraisal generally qualitative, subjective, and documented in free text clinical notes rather than as structured data. Previous attempts to analyse EHRs have shown to lead to low sensitivity regarding identification of suicidal behaviour when relying on codified data points such as International Classification of Diseases (ICD) billing codes^6^.

Natural language processing (NLP) and its subdiscipline of Information Extraction (IE) are commonly employed and tested within clinical records in the fields of, for instance, oncology and radiology^7,8^ to process large quantities of unstructured (human authored) text and return structured information about its meaning. However, there has been little application of NLP techniques to mental healthcare data, despite the volumes of text-based information contained, and even less for suicide risk assessment. Few quantitative studies have been published on analysing English text from medical records in the psychiatric field. Irving et al.^3^ successfully used NLP tools to extract symptom and substance use data from free text recorded by clinicians to enhance the prognostic accuracy of a psychosis risk calculator. Other implementations of NLP in psychiatry include its use to classify patients with first episode psychosis and distinguish them from healthy controls based on their speech documented in interview transcripts^9^, to extract information on risk factors for hospital readmission^4^, to identify constellations of symptoms in patients with schizophrenia spectrum disorders and other severe mental illness based on clinical records^10–12^.

Some novel approaches have attempted to use NLP to determine the possibility of suicide attempts within specific patient subgroups such as US Army veterans with diagnosed post-traumatic stress disorder (PTSD)^13^, adolescents with Autism Spectrum Disorders (ASD)^14^, pregnant women^6^ or individuals without any previous suicide attempts^15^. For instance, Tsui et al.^15^ employed the clinical Text Analysis and Knowledge Extraction System (cTAKES™) to extract information from EHRs without pre-processing followed by the application of four different widely used Machine Learning (ML) methods on the data. Utilising these methods, they achieved varying results for the different ML techniques. Moreover, Senior et al.^5^ used free-text entries in medical records to assess the presence of predictors of suicide risk. In order to achieve this, through annotation, they trained a named entity recognition tool using a neural network algorithm.

From the above, it is evident that NLP can be used in order to analyse unstructured clinical documentation, but not without challenges. Conventionally, NLP is only functional after having defined rules. While this might be achievable with regards to the analysis of structured data, once dealing with unstructured input it is a challenge to list all such rules manually beforehand.

To overcome this hurdle, machine learning and more recently deep learning (DL) models have been used to study mental health disorders^16^. DL refers to a method for analysing large amounts of data through various layers of representations meant to increase accuracy. One example for this is the technique of long short term memory (LSTM)^17^ with its recurrent feedback cycles which have been applied to handwriting recognition^18^, sequence modelling^19^ and as well as widely utilised in connection with NLP^20–22^.

Moreover, LSTMs have been successfully implemented to predict psychiatric outcomes in recent studies. For instance, Pham et al.^23^ have applied an LSTM-based DL model to predict the future outcomes of depressive episodes based on patients’ long-term health state trajectories. Further, Song et al.^24^ used a Context-LSTM-convolutional neural networks (C-LSTM-CNN) model to improve the predictive accuracy of suicide risk assessment previously established by Downs et al.^14^ in adolescents with ASD.

As DL techniques continue to be refined and improved upon, it will be possible to help mental health professionals assess suicidality in a more standardized manner, identify the corresponding risk factors at an earlier stage -when interventions may be more effective-, and personalize interventions based on an individual’s unique characteristics. A quantitative approach (as opposed to the usually qualitative approach in current clinical practice) will add a considerable amount of value due to its fundamentally different nature: While qualitative methods allow for an interactive approach with the goal of finding explanations for and interpretations of observed phenomena, quantitative methods introduce less bias as there is independence between researchers and participants, while at the same time gathering data that can be generalised to a larger population, and used for prediction of future occurrences^25^.

Research so far has only explored the potential of NLP application for suicide prediction on a limited scale with varying accuracy, often focussing on single specific cohorts, or using highly specific terms and concepts unique to the corresponding group. Further, NLP using the German language is still underrepresented in these medical use cases and is mostly applied to structured data or standardised speech. A German NLP application in the field of psychiatry and specifically for use in suicide prediction remains to be seen.

The present study aims to assess suicide risk during inpatient treatment in a manner quantifiable and applicable to an everyday clinical setting, by using NLP and DL to extract pertinent information from German-language medical records. We will not limit our approach to a specific subgroup but will include all reported cases of suicide and suicide attempt of patients admitted to two university psychiatric departments in the past 8 years, to develop a tool that can be implemented more universally. In this study, we focused on prediction of risk factors on mention level, certain rules for deduction similar to those of Song et al.^24^ or Downs et al.^14^ are necessary to extrapolate the data to a patient level.

## Methods

### A. Study Population and Data Privacy

Participants who died by suicide and those with a previous suicide attempt will be collected from the EHR system of the Department of Adult Psychiatry and Psychotherapy and the Department of Psychosomatics and Psychotherapy of the Centre of Integrative Psychiatry (ZIP), Campus Lübeck. Suicide attempts (completed or not) will be identified by filtering through a manual and computer assisted search for the following ICD-10 codes at discharge described as relevant by Hedegaard et al.^26^ and adapted by members of the research group: R45.8, T36-T65, T71, T14.91, X60-X84, Z91.8.

Patient records will be included in the analysis as part of the study cohort if they meet the following inclusion criteria:

1. Admission into one of the inpatient or day care units of the Departments of Adult Psychiatry, or Psychotherapy or Psychosomatics and Psychotherapy, of the Centre of Integrative Psychiatry, Campus Lübeck.
2. Admission date from 2013 to 2021 as electronic health records at these departments were first applied in 2013.
3. Completed suicide while receiving inpatient or day care treatment documented in the electronic health records of the departments.

An external validation dataset will be collected using the same filtering criteria from the Department of Adult Psychiatry and Psychotherapy and the Department of Psychosomatics and Psychotherapy of the Centre of Integrative Psychiatry, Campus Kiel.

To comply with national data protection laws and local ethics regulations, patients’ health records will be anonymised at the source to redact any personal, immediately identifying data, thus improving data privacy and patient safety. The hospital’s IT department will anonymize the data before the transfer.

The name and the date of birth will not be available, instead the age will be considered as a variable. The total number of cases will only be known after data collection and filtering through the methods explained above. The study procedures were approved by the ethics committee of the University of Lübeck on September 28^th^, 2021 (reference: 21-256).

### B. Data collection

Patients’ health records are kept in a digital patient data management system combining health care, organizational and administrative documentation in an electronic format. For this study, we will focus on clinical reports, especially discharge letters, everyday clinical notes and medical and psychiatric history notes recorded on admission. Clinical reports for the study will be collected as pdf documents and anonymized at the source. Following this step, the text will be extracted from the pdf documents to be fed into the workflow explained below. All clinical reports by mental health care professionals uploaded on the day of admission, as well as the 10 day-period before the attempted or completed suicide will be evaluated. For control patients, documentation of the 10 days before the day of discharge will be evaluated. Should a hospitalisation have lasted less than 10 days, all available data from admission to discharge will be considered.

### C. Workflow

A corpus will be created from unstructured EHR text that will be manually annotated for suicide risk factors. A list of concepts has been gathered and is continuously being added to by the research team based on professional experience and the concepts used by Senior et al.^5^ and Zhong et al.^6^. The annotated corpus will be used to train NLP applications to extract such suicide risk factors from the EHR and validated for use in suicide prediction tasks. The full list will be attached to the final study in the appendix.

The structure is based on the C-LSTM-CNN model described by Song et al.^24,27^ and uses three inputs as described by these authors: 1. the focus sentence - the sentence meant to be classified; 2. left context - all document text to the left of the focus sentence; 3. right context - all document text to the right of the focus sentence. The fixed size ordinally-forgetting encoding (FOFE) is described below. The full workflow is presented in **Figures 1 and 2**:

**Figure 1:**
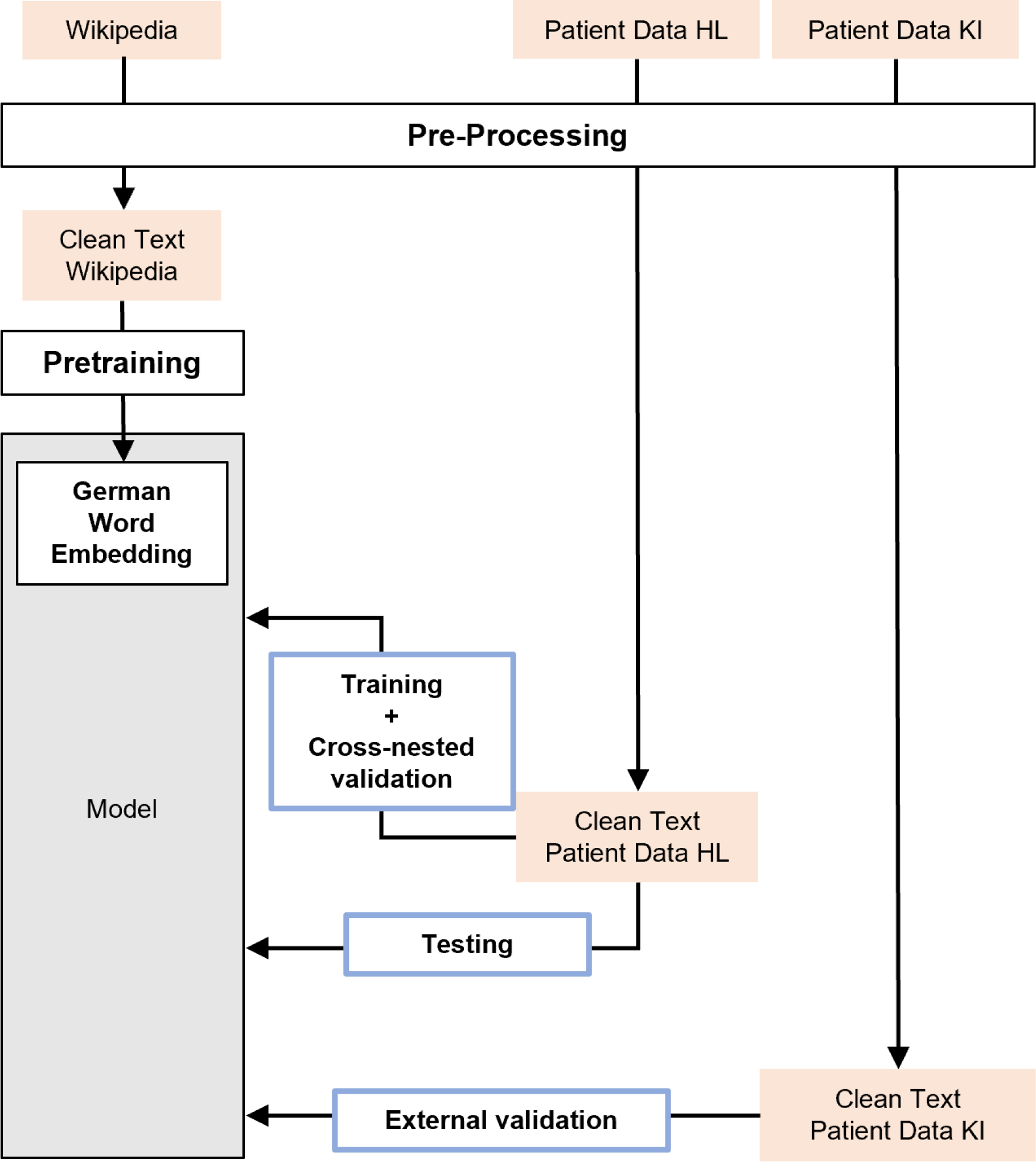
Overview Pre-Processing, Training and Evaluation (HL = Lübeck, KI = Kiel)

**Figure 2:**
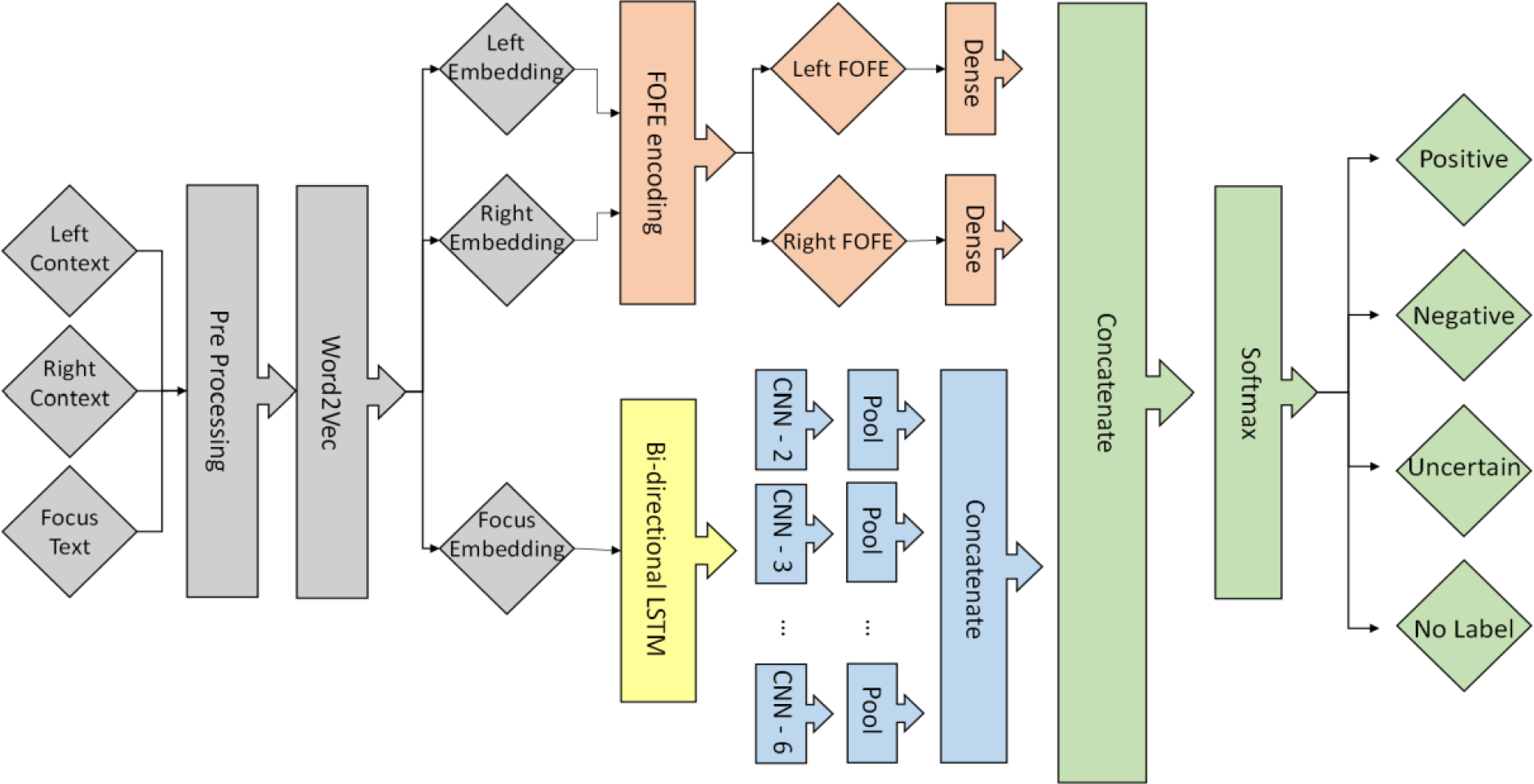
Modelling workflow chart based on *Song et al*.^24^

The full number of documents for the prediction of a) suicidal behavior is 1,395 and b) completed suicide is 458. We will be using a maximum of 400 documents for the annotation process for case a) and 200 documents for case b); an amount comparable to that used by Senior et al.^5^ and Zhong et al.^6^ in their respective approaches and sufficient to allow for meaningful comparisons between models. These number of documents will make up the model training set for the 2 cases. For the case a) the documents from case b) will be pooled together.

Within this model training set we will employ a 10×10 repeated nested cross-validation scheme composed of 10 folds in the inner as well as 10 folds in the outer cycle depicted in **Figure 3**. The documents in question make up the dataset and will be split into these 10 folds of which one will remain the hold-out dataset while the other 9 are used for training further referred to as the training set. Similarly, after dividing said training set into 10 folds, 9 of them will be used for training while the spare will be used for validation consisting of model parameter selection as well as determination of the winner model, the model with the best-balanced accuracy during validation. Following this, the hold-out dataset will be fed into the network. This process will be applied 10 times, each time different through shuffling the primary dataset resulting in 10,000 models. After completion, we will determine the average accuracy achieved in the hold-out dataset across the 10 times repeated 10×10 nested cross-validation. Further external validation will be done using the data collected from the Campus Kiel.

**Figure 3:**
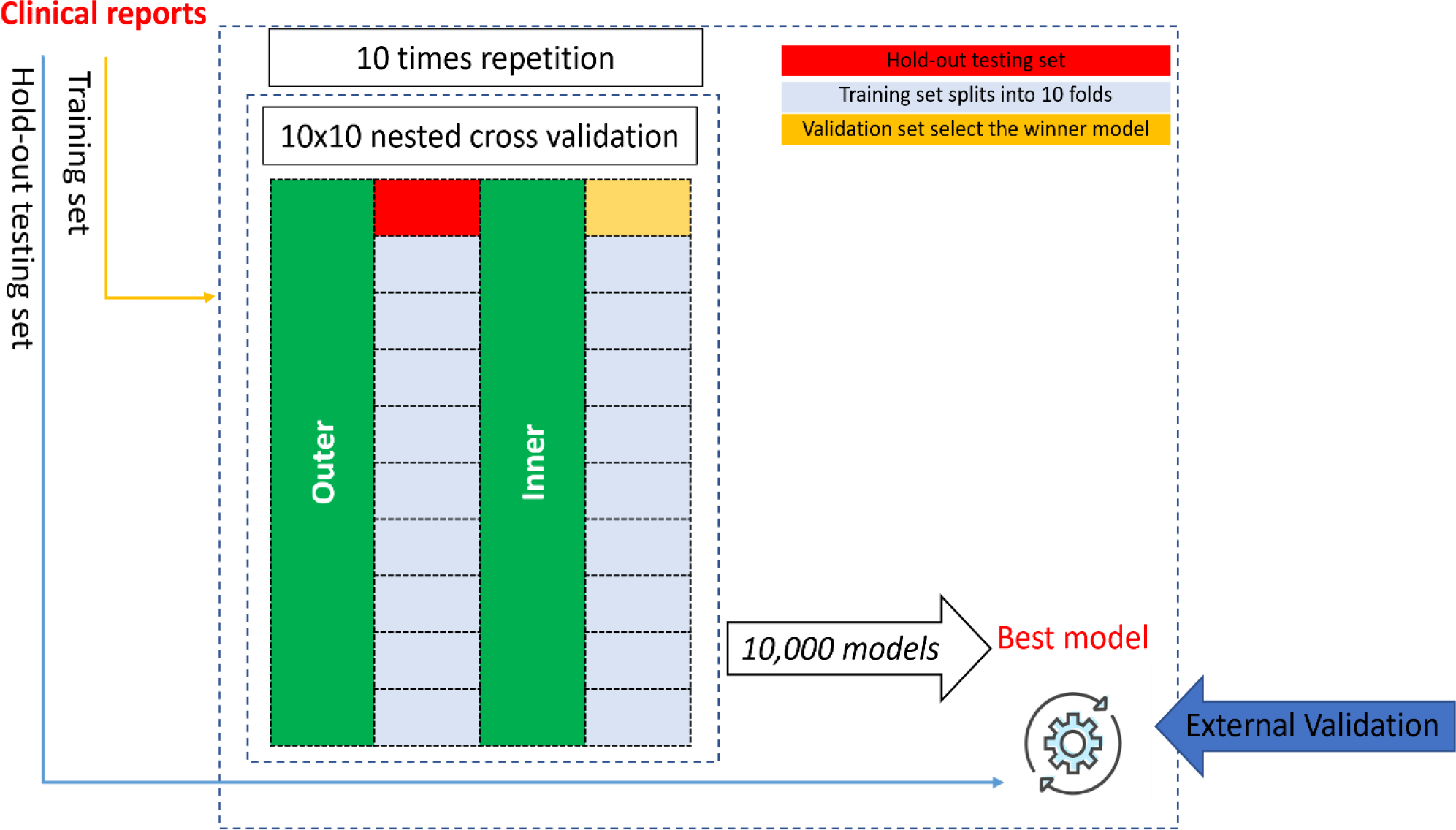
Nested cross-validation scheme based on *source*

#### Pre-Processing

To be able to process the acquired medical documents tokenization is necessary. In this case word tokenisation is applied by using the Natural Language Toolkit (NLTK) to separate each individual word^28^. These are then used as input for the word embedding layer. Further, characters or words unimportant for the analysis will be removed including punctuation as well as certain common words (e.g. “Dr.”, “Arzt/Ärztin”, “Hr.”, “Fr.”), and umlauts will be replaced by their international digraph representation (combination of codes adapted from methods found <u>here</u> and <u>here</u>). The pre-processing steps are presented in **Figure 4**.

**Figure 4:**
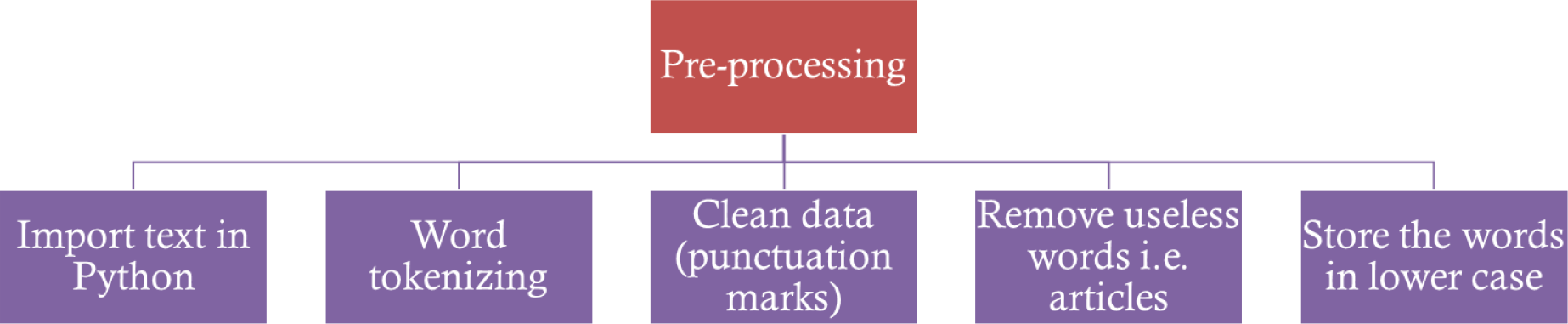
Pre-Processing components

#### Document annotation

The model training set text will then be annotated by one medical student in accordance with the full list of concepts (see appendix) supervised by one mental health professional with clinical experience in psychiatry.

Sentences that include mentions of one or more concepts will be marked as sentences of interest. Further information will be given by labelling a mention as Suicide related (SR) positive or SR negative,in the beginning and two more labels will be added later; uncertain or no label. Sentence level labelling will be derived from the combination of mention levels considering the following scenarios:

- Only one mention label: sentence will be assigned same label as mention
- Multiple mentions all with the same label: sentence will be assigned same label as mentions
- Multiple mentions of different labels incl. ≥1 SR positive mention: sentence will be given the SR positive label
- No SR positive mentions but ≥1 uncertain mention label: sentence will be given the uncertain label
- None of the above: sentence will be given no label

#### Word embedding: Word2Vec and numeric representation

For DL to process the free text documents taken from the EHRs, the texts have to be converted into structured data representations. Specifically, the starting point of the proposed algorithm will be Word2Vec^29^ word embeddings *e*_*u*_ of each word, grouped together to a sentence *S* of length *U* with an index *i* indicating the position of the sentence in question in a text with *I* number of sentences in total: *S*_*i*_ = (*e*_1_, *e*_2_, …, *e*_*U*_), *i* = 1,.., *I*. Word2Vec has been implemented in the TensorFlow tool for the English language (https://www.tensorflow.org/tutorials/text/word2vec). We will train the Word2Vec using German free text, after pre-processing it as described in **Figure 3**. The resulting embedding algorithm will be evaluated using text files containing German grammatical examples and validated using a text corpus consisting of German news articles. The evaluated model will be applied on our corpus consisting from the pre-processed pdf documents received from our clinic.

#### LSTM/CNN

Following the above pre-processing steps bi-directional Long Short-Term Memory (LSTM) and Convolutional Neural Networks (CNN) will be applied to the sentence embeddings. The intention is to take into consideration connections of words within the same sentence during encoding and have the model recognize particular features. The ideal number of hidden layers of the LSTM or kernel size of the CNN-layers cannot be predetermined as their efficacy will only be measurable after implementation. However, we will start by using the parameters mentioned in Song et al.^24^ and will therefore be employing 64 hidden LSTM-layers in addition to CNNs with kernel sizes ranging from 2 to 6, each in turn using 64 features.

#### FOFE

Our approach is a form of DL called a feedforward neural network (FNN) that only allows for fixed size input, while the previously mentioned context consists of a fluctuating number of sentences of varying length. To overcome this challenge, we will employ the FOFE method as developed by Zhang et al.^30^ and adapted by Song et al.^24,27^. This procedure employs a predefined forgetting factor and considers the word order within a sentence to uniquely, for the most part, generate a fixed size representation in code. Following the conversion of words into word embeddings *e*_*u*_, these will be the used in place of the 1- of-K representation originally described for the FOFE model^30^. In detail, starting from *u* = 1, …, *U*, with *U* being the number of words in a specific sentence *S*_*i*_ = {*e*_1_, *e*_2_, …, *e*_*U*_}, *i* = 1, …, *I*, FOFE will encode partial word representation sequences *e*_*u*_ for each word as shown in **Figure 5** based on the recursive formula (1).

**Figure 5:**
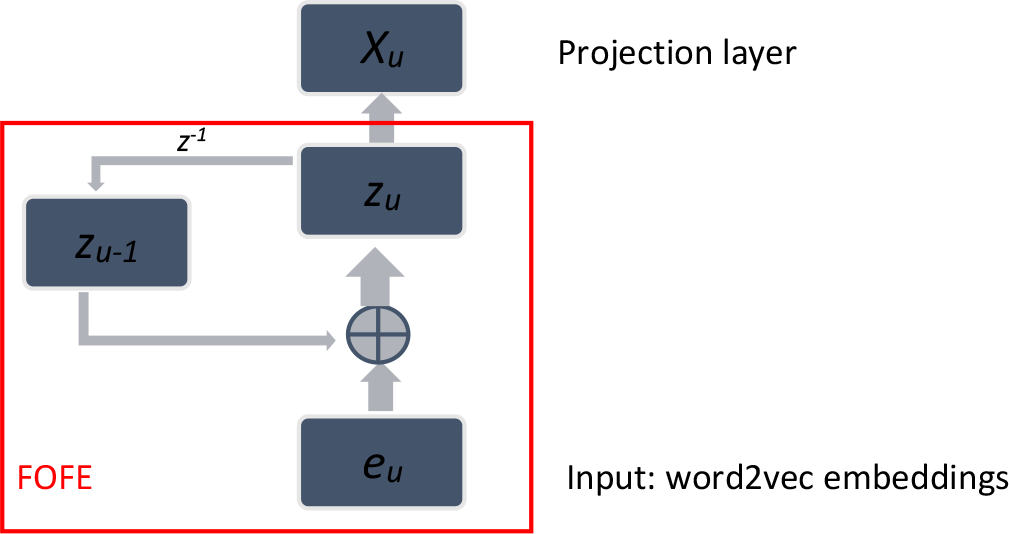
FOFE model based on work by *Zhang et al*.^30^

In order to be able to condense all the information of the left and right context, a hierarchical two-step FOFE process will be implemented, starting with each sentence being encoded separately (*z*_*sent*_*i*_, *i* = 1,.., *I*). For this embedding, a slowly-decreasing variable *α*_*sent*_, 0 < *α*_*sent*_ < 1 will be used which leads to words within this sentence only being slightly differently weighted in proportion to their distance to the focus sentence, as expressed in formula (2).

In the next step, when considering the full context on either side, a rapidly decreasing *α*_*cont*_ ensures that sentences in close proximity to the focus sentence are awarded high importance within the algorithm with those furthest away are awarded less importance. The formulas for the left and right context embeddings are expressed in (3) and (4) below.

##### FOFE formulas based on findings by Song et al.^24,27^

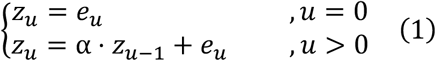

where *z*_*u*_ denotes the FOFE code for the partial sequence up to *e*_*u*_.

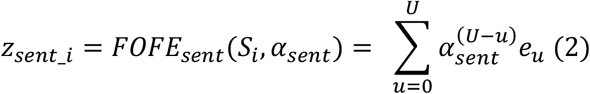

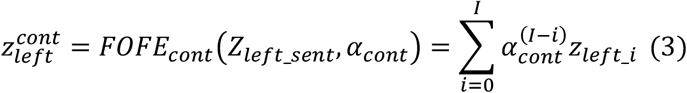

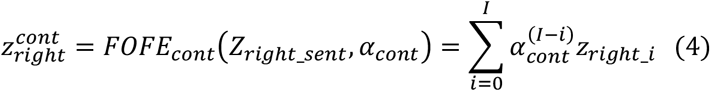

#### Evaluation

For evaluation, average accuracy, recall, precision, F-measure and AUC will be taken into consideration for each of the labelling categories (suicidality risk positive, suicidality risk negative, uncertain, no label) in the test set.

Its results will be compared with scores achieved by applying NegTool as one alternative representing only the NLP approach and separately applying Support vector machine (SVM) as a Machine Learning (ML) alternative as done in Song et al.^24^

#### Preliminary Results for the completed suicide dataset

We identified 10 patients who committed suicide during their in-patient psychiatric hospitalisation between 2011 and 2022. The decision support system on sentence level was trained and validated with 259 positively labelled sentences and 5,016 negatively labelled sentences. Preliminary results showed a mean validation accuracy of 76.98% and precision of 63.49% on sentence level across both labels. Mean precision for positive labels was 40.98% while mean precision for negative labels was 86.0%.

## Discussion

Through further training using more cases’ documentation as well as expanding labelling criteria the algorithm can be improved significantly in the future. Prospectively, the sentence labels generated by the algorithm can be used to extrapolate the information to a document or patient level and thus give a prediction on the suicidality of patients as a whole. This study will offer promising ways for automated early detection of suicide and therefore holds opportunities for mental health care.

## Data Availability

All data produced in the present study are available upon reasonable request to the authors

## Acknowledgements

*The preliminary results were accepted for poster presentation at SIRS conference 2023 and the International Schizophrenia Symposium 2023 in Giessen*.

### Appendix

#### Full list of concepts

This list was formed based on professional experience of members of the research team as well as concepts used in Senior et al.^5^ and Zhong et al.^6^.

|  |  |
| --- | --- |
| *Age* | *Length of first inpatient stay* |
| *Sex* | *Presence of comorbid schizophrenia and depression* |
| *Previous suicide attempts* | *Highest education* |
| *Previous drug abuse* | *Receipt of welfare or disability benefit* |
| *Previous alcohol abuse* | *Previous conviction of a violent offence* |
| *Previous self-harm* | *Parental psychiatric hospitalization* |
| *Recent antipsychotic treatment* | *Parental drug or alcohol use disorder* |
| *Recent antidepressant treatment* | *Parental suicide* |
| *Feeling hopeless* | *Isolation* |
| *Feeling relief* | *Distractibility* |
| *Tired* | *Childhood adversity* |
| *Love* | *Adult sexual abuse* |
| *Feeling empty* | *Severe depression* |
| *Feeling content* | *Substance abuse problem* |
| *Low self-esteem* | *Personality disorder* |
| *Impulsive character* | *Psychotic disorders* |
| *Seizures* | *Anxiety disorders* |
| *Wound and injury* | *Abortion* |
| *Forced admission/hospitalization* |  |

## References

1. World Health Organization. Suicide. https://www.who.int/news-room/fact-sheets/detail/suicide (2021).

2. Statistisches Bundesamt. Suizide. https://www.destatis.de/DE/Themen/Gesellschaft-Umwelt/Gesundheit/Todesursachen/Tabellen/suizide.html (2021).

3. Irving, J. et al. Using Natural Language Processing on Electronic Health Records to Enhance Detection and Prediction of Psychosis Risk. Schizophr. Bull. 47, 405–414 (2021).

4. Holderness, E. et al. Analysis of risk factor domains in psychosis patient health records. J. Biomed. Semant. 10, 19 (2019).

5. Senior, M. et al. Identifying Predictors of Suicide in Severe Mental Illness: A Feasibility Study of a Clinical Prediction Rule (Oxford Mental Illness and Suicide Tool or OxMIS). Front. Psychiatry 11, 11 (2020).

6. Zhong, Q.-Y. et al. Use of natural language processing in electronic medical records to identify pregnant women with suicidal behavior: towards a solution to the complex classification problem. Eur. J. Epidemiol. 34, 153–162 (2019).

7. Bitterman, D. S., Miller, T. A., Mak, R. H. & Savova, G. K. Clinical Natural Language Processing for Radiation Oncology: A Review and Practical Primer. Int. J. Radiat. Oncol. Biol. Phys. 110, 641–655 (2021).

8. Pons, E., Braun, L. M. M., Hunink, M. G. M. & Kors, J. A. Natural Language Processing in Radiology: A Systematic Review. Radiology 279, 329–343 (2016).

9. Si, D., Cheng, S. C., Xing, R., Liu, C. & Wu, H. Y. Scaling up Prediction of Psychosis by Natural Language Processing. in 2019 IEEE 31st International Conference on Tools with Artificial Intelligence (ICTAI) 339–347 (2019). doi:10.1109/ICTAI.2019.00055.

10. Jackson, R. G. et al. Natural language processing to extract symptoms of severe mental illness from clinical text: the Clinical Record Interactive Search Comprehensive Data Extraction (CRIS-CODE) project. Open Access 10 (2017).

11. Liu, Q. et al. Symptom-based patient stratification in mental illness using clinical notes. J. Biomed. Inform. 98, 103274 (2019).

12. Su, C., Xu, Z., Pathak, J. & Wang, F. Deep learning in mental health outcome research: a scoping review. Transl. Psychiatry 10, 1–26 (2020).

13. Levis, M., Leonard Westgate, C., Gui, J., Watts, B. V. & Shiner, B. Natural language processing of clinical mental health notes may add predictive value to existing suicide risk models. Psychol. Med. 1–10 (2020) doi:10.1017/S0033291720000173.

14. Downs, J. et al. Detection of Suicidality in Adolescents with Autism Spectrum Disorders: Developing a Natural Language Processing Approach for Use in Electronic Health Records. AMIA Annu. Symp. Proc. AMIA Symp. 2017, 641–649 (2017).

15. Tsui, F. R. et al. Natural language processing and machine learning of electronic health records for prediction of first-time suicide attempts. JAMIA Open 4, ooab011 (2021).

16. Lin, E. et al. A Deep Learning Approach for Predicting Antidepressant Response in Major Depression Using Clinical and Genetic Biomarkers. Front. Psychiatry 9, (2018).

17. Hochreiter, S. & Schmidhuber, J. Long Short-Term Memory. Neural Comput. 9, 1735–1780 (1997).

18. Graves, A. et al. A Novel Connectionist System for Unconstrained Handwriting Recognition. IEEE Trans. Pattern Anal. Mach. Intell. 31, 855–868 (2009).

19. Graves, A. Generating Sequences With Recurrent Neural Networks. ArXiv13080850 Cs (2014).

20. Maragatham, G. & Devi, S. LSTM Model for Prediction of Heart Failure in Big Data. J. Med. Syst. 43, 1–13 (2019).

21. Ghosh, S. et al. Contextual LSTM (CLSTM) models for Large scale NLP tasks. ArXiv160206291 Cs (2016).

22. Xu, K., Zhou, Z., Hao, T. & Liu, W. A Bidirectional LSTM and Conditional Random Fields Approach to Medical Named Entity Recognition. in Proceedings of the International Conference on Advanced Intelligent Systems and Informatics 2017 355–365 (Springer, Cham, 2017). doi:10.1007/978-3-319-64861-3_33.

23. Pham, T., Tran, T., Phung, D. & Venkatesh, S. Predicting healthcare trajectories from medical records: A deep learning approach. J. Biomed. Inform. 69, 218–229 (2017).

24. Song, X. et al. Using Deep Neural Networks with Intra- and Inter-Sentence Context to Classify Suicidal Behaviour. in Proceedings of the 12th Language Resources and Evaluation Conference 1303–1310 (European Language Resources Association, 2020).

25. Yilmaz, K. Comparison of Quantitative and Qualitative Research Traditions: epistemological, theoretical, and methodological differences. Eur. J. Educ. 48, 311–325 (2013).

26. Hedegaard, H. et al. Issues in Developing a Surveillance Case Definition for Nonfatal Suicide Attempt and Intentional Self-harm Using International Classification of Diseases, Tenth Revision, Clinical Modification (ICD-10-CM) Coded Data. Natl. Health Stat. Rep. 19 (2018).

27. Song, X., Petrak, J. & Roberts, A. A Deep Neural Network Sentence Level Classification Method with Context Information. in Proceedings of the 2018 Conference on Empirical Methods in Natural Language Processing 900–904 (Association for Computational Linguistics, 2018). doi:10.18653/v1/D18-1107.

28. Loper, E. & Bird, S. NLTK: The Natural Language Toolkit. arXiv:cs/0205028 (2002).

29. Mikolov, T., Chen, K., Corrado, G. & Dean, J. Efficient Estimation of Word Representations in Vector Space. ArXiv13013781 Cs (2013).

30. Zhang, S., Jiang, H., Xu, M., Hou, J. & Dai, L. The Fixed-Size Ordinally-Forgetting Encoding Method for Neural Network Language Models. in Proceedings of the 53rd Annual Meeting of the Association for Computational Linguistics and the 7th International Joint Conference on Natural Language Processing (Volume 2: Short Papers) 495–500 (Association for Computational Linguistics, 2015). doi:10.3115/v1/P15-2081.

31. Cirillo, D. et al. Sex and gender differences and biases in artificial intelligence for biomedicine and healthcare. Npj Digit. Med. 3, 81 (2020).

32. Garg, N., Schiebinger, L., Jurafsky, D. & Zou, J. Word embeddings quantify 100 years of gender and ethnic stereotypes. Proc. Natl. Acad. Sci. 115, E3635–E3644 (2018).

